# Hereditary breast cancer next-generation sequencing (NGS) panel evaluation in the south region of Brazil: a novel *BRCA2* candidate pathogenic variant is reported

**DOI:** 10.1101/2024.02.08.24302195

**Authors:** Cesar Augusto B. Duarte, Carlos Alberto dos Santos, Cristine Domingues D. de Oliveira, Cleverton César Spautz, Laura Masami Sumita, Sueli Massumi Nakatani

**Affiliations:** Research and Development Division, Genoprimer Diagnóstico Molecular, Curitiba, Paraná, Brazil; Clinimol Diagnóstico Molecular, Molecular Biology Clinical Laboratory, São Paulo, São Paulo, Brazil; Hospital Nossa Senhora das Graças, Department of Surgery, Curitiba, Paraná, Brazil

## Abstract

In this article, we delineate a loosely selected cohort comprising patients with a history of early-onset breast cancer and/or a familial occurrence of cancer. The aim of this study was to gain insights into the presence of breast cancer-related gene variants in a population from a micro-region in southern Brazil, specifically the Metropolitan Region of Curitiba. This area exhibits a highly genetically mixed population, mirroring the general characteristics of the Brazilian people. Comprehensive next-generation sequencing (NGS) multigene panel testing was conducted, involving the evaluation of twelve patients. Two pathogenic variants and one candidate pathogenic variant were identified: *BRCA2*:c.8878C>T, p.Gln2960Ter; *CHEK2*:c.1100delAG>A, p.Thr367Metfs*15 and *BRCA2*:c.3482dupG>GA, p.Asp1161Glufs*3, a novel variant, previously unpublished, is reported.

**Author Summary:** Breast cancer stands as one of the most prevalent cancers globally, affecting both genders, although it is much more common in women. Predominantly sporadic, around 5% to 10% of cases are attributed to hereditary factors, linked to specific gene mutations passed down through generations. Beyond *BRCA1* and *BRCA2*, there is a growing understanding that breast cancer risk is influenced by a range of genes. In the pursuit of understanding and mitigating breast cancer risks, genetic testing plays a pivotal role. These tests draw upon extensive databases, repositories of genetic information, to decipher individual variations. However, the lack of population diversity representation in genetic research databases is a global concern, and Latin America is no exception, presenting challenges in ensuring inclusivity and relevance in genetic research and healthcare applications. Addressing this gap, our study focused on a population in the Metropolitan Region of Curitiba, Brazil, sought to uncover breast cancer-related genetic variants. Despite the modest cohort, the study identified two pathogenic and one novel candidate pathogenic variants. While a small contribution, this research endeavors to enrich the collective knowledge base surrounding breast cancer, illustrating the ongoing efforts to comprehend and address the complexities of this prevalent disease.

## Introduction

In Brazil, breast cancer ranks as the most prevalent cancer in women across all regions, second only to non-melanoma skin cancer. The estimated number of new cases in 2024 is approximately 70,000[1]. While hereditary factors contribute to less than 10% of breast cancer cases, identifying carriers of pathogenic variants associated with increased cancer risk remains a potentially cost-effective healthcare strategy, given the high annual case volume and improved prognosis with early detection[2].

The past decade has witnessed widespread adoption of next-generation sequencing (NGS) platforms and advanced bioinformatic tools, enabling the identification of numerous pathogenic variants in individuals with clinical presentations of hereditary breast and ovarian cancer (HBOC). The interpretation of sequence variants, guided by the American College of Medical Genetics and Genomics (ACMG) and the Association for Molecular Pathology (AMP) guidelines, relies on comprehensive databases and published literature[3].

This study emphasizes the significance of reporting pathogenic/likely pathogenic variants, including conflicting interpretations, to stimulate further discussions. Acknowledging the bias in genetic association studies towards European populations[4], our modest contribution aims to broaden the community’s knowledge. Despite the limited sample size impacting the feasibility of future screening strategies, the study seeks to identify SNPs/Indels variants associated with breast cancer, offering potential insights for future research.

While efforts have been made to advance the diagnosis and management of HBOC in Brazil[5], it’s noteworthy that a recent comprehensive review of Brazilian germline mutations in *BRCA1* and *BRCA2* did not include patients from the specific region addressed in our study[6]. Consequently, our findings aim to fill this gap, providing accurate and actionable information to contribute meaningfully to the field.

## Results

The standards and guidelines for interpreting sequence variants, as outlined by the ACMG/AMP, categorize variants into pathogenic, likely pathogenic, uncertain significance, likely benign, or benign. In essence, this guideline employs a set of criteria assigned to each evaluated variant with the intention of classifying it as pathogenic or benign. The summation of these criteria values leads to the classification of a variant into one of the five tiers mentioned above. Twelve variants were identified and classified as either pathogenic (P)/likely pathogenic (LP) or variants of uncertain significance (VUS), and these are reported here.

Among the 12 patients, three P/LP variants were identified, constituting 25% of the cases. Most of the listed variants were detected in all three raw data sets generated from the library preparation methods used in this evaluation (see Supporting information, S1 Table). The three variants classified as either P/LP shared the commonality of a null variant effect in a gene where the loss of function is a known mechanism of disease *(BRCA2*:c.3482dup, *CHEK2*: c.1100del and *BRCA2*:c.8878C>T). All of them met the very rare frequency criterion in population databases; variant c.3482dup was not found in any database, and to the best of our knowledge, it has never been described before.

Variant *PMS2*:c.2186_2187delTGA>T, was the only variant with a null variant effect in a gene where the loss of function is a known mechanism of disease, that was not classified as pathogenic/likely pathogenic due to the absence of additional criteria for such. Pertinent considerations about this variant will be addressed later in the discussion section.

Other variants described here were also classified as variants of unknown significance. These variants shared a moderate level of evidence of pathogenicity, based on their absence or extremely low frequency in all databases. Additionally, they exhibited other attributes such as in-silico predictions, effects on protein, and functional data that were insufficient for classification as either pathogenic or benign.

The identified variants and the respective adopted criteria are summarized in Tables 1 and 2, with references to ClinVar[7] and Single Nucleotide Polymorphism database(dbSNP)[8].

**Table 1.**
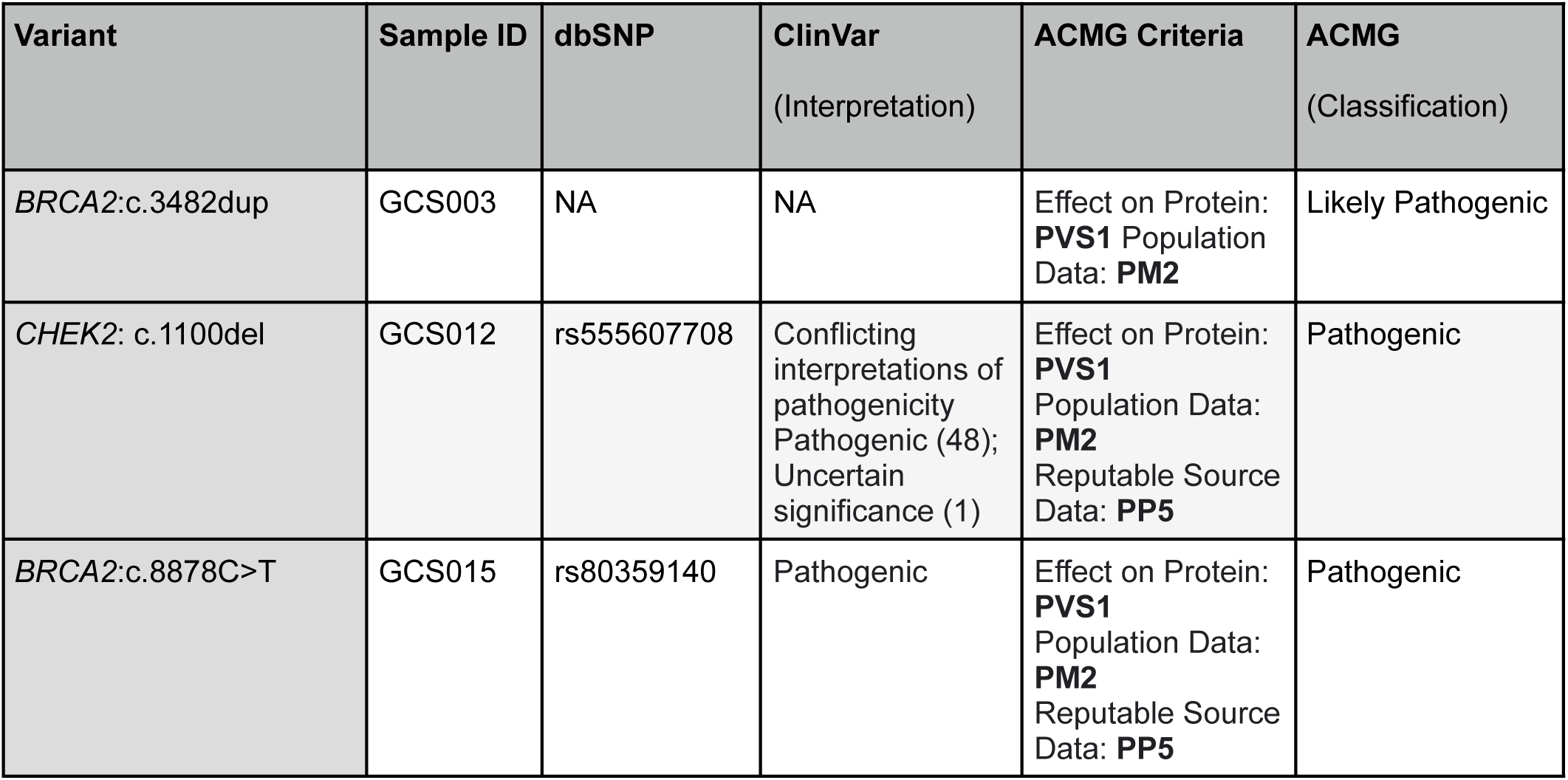
Pathogenic/likely pathogenic variants.

**Table 2.**
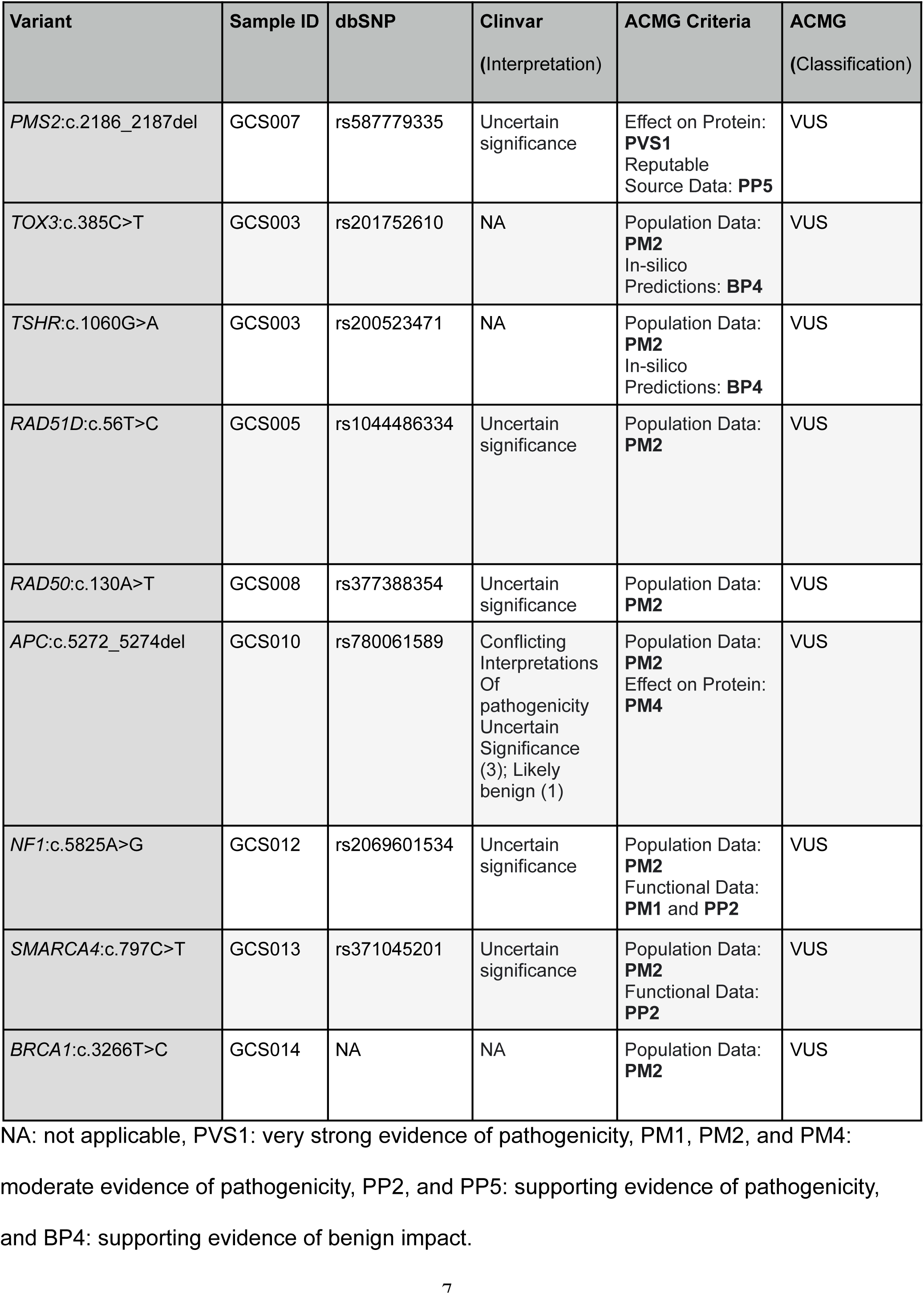
Variants of uncertain significance.

| Variant | Sample ID | dbSNP | Clinvar<br>(Interpretation) | ACMG Criteria | ACMG<br>(Classification) |
| --- | --- | --- | --- | --- | --- |
| *PMS2*:c.2186_2187del | GCS007 | rs587779335 | Uncertain<br>significance | Effect on Protein:<br>**PVS1**<br>Reputable<br>Source Data: **PP5** | VUS |
| *TOX3*:c.385C>T | GCS003 | rs201752610 | NA | Population Data:<br>**PM2**<br>In-silico<br>Predictions: **BP4** | VUS |
| *TSHR*:c.1060G>A | GCS003 | rs200523471 | NA | Population Data:<br>**PM2**<br>In-silico<br>Predictions: **BP4** | VUS |
| *RAD51D*:c.56T>C | GCS005 | rs1044486334 | Uncertain<br>significance | Population Data:<br>**PM2** | VUS |
| *RAD50*:c.130A>T | GCS008 | rs377388354 | Uncertain<br>significance | Population Data:<br>**PM2** | VUS |
| *APC*:c.5272_5274del | GCS010 | rs780061589 | Conflicting<br>Interpretations<br>Of<br>pathogenicity<br>Uncertain<br>Significance<br>(3); Likely<br>benign (1) | Population Data:<br>**PM2**<br>Effect on Protein:<br>**PM4** | VUS |
| *NF1*:c.5825A>G | GCS012 | rs2069601534 | Uncertain<br>significance | Population Data:<br>**PM2**<br>Functional Data:<br>**PM1** and **PP2** | VUS |
| *SMARCA4*:c.797C>T | GCS013 | rs371045201 | Uncertain<br>significance | Population Data:<br>**PM2**<br>Functional Data:<br>**PP2** | VUS |
| *BRCA1*:c.3266T>C | GCS014 | NA | NA | Population Data:<br>**PM2** | VUS |

NA: not applicable, PVS1: very strong evidence of pathogenicity, PM1, PM2, and PM4:
moderate evidence of pathogenicity, PP2, and PP5: supporting evidence of pathogenicity,
and BP4: supporting evidence of benign impact.

## Discussion

Although our NGS panel was comprehensive, it is crucial to note that the risk posed by pathogenic variants found in these genes is not evenly distributed. Gene risk stratification for HBOC has been proposed elsewhere[9]. For example, *BRCA1* and *BRCA2* gene mutations are responsible for the majority of HBOC cases. However, the role of *MSH6* and *PMS2* is less clear and has been a matter of dispute[10,11]. Ordering physicians and patients alike must be aware of the implications associated with the selected gene panel scope, including risks and actionability. With the increase in the volume of generated sequencing data over the last decades, the challenge of accurately verifying the consequences of a diverse array of variants has grown. Even with a limited gene panel sequencing, uncertainties may arise. For the two variants classified as pathogenic by ACMG criteria, the ClinVar database indicates that *BRCA2*:c.8878C>T has been reviewed by an expert panel and classified as pathogenic. *CHEK2*:c.1100del is listed under “conflicting interpretations of pathogenicity” in that database, although there is overwhelming reporting of pathogenicity with 48 instances, and uncertain significance reported in one instance. We understand that both variants should be reported as pathogenic. *BRCA2*:c.3482dup, classified as likely pathogenic (ACMG), has not been reported in any database to the best of our knowledge, including BRCA Exchange[12], Genome Aggregation Database (gnomAD)[13], and ClinVar. Incidentally, this variant is located in the *BRCA2* ovarian cancer cluster regions (OCCRs), where a small but statistically significant difference in the mean age at breast cancer diagnosis was found[14]. The mean age was greater for mutations in OCCR compared to mutations not in OCCR. This information may be relevant to the carrier of this variant, who, in our study, is described as having relatives diagnosed with breast cancer but no personal history of cancer in her 50s.

The frameshift-caused truncated protein is rather similar to two other known pathogenic variants listed in gnomAD: variants *BRCA2*:c.3481_3482dup chr13-32337832 C>CAG p.Asp1161Glufs8 and *BRCA2*:c.3487del chr13-32337841 TG>T p.Asp1163Ilefs5, which would cause truncated proteins slightly larger than *BRCA2*:c.3482dup. Consequently, we conclude that this variant should be classified as pathogenic instead of likely pathogenic. Regarding the remaining reported variants classified as VUS, we were unable to gather evidence to reclassify them as either pathogenic or benign. However, *PMS2*:c.2186_2187del, a null variant in a gene where the loss of function is a known mechanism of disease, exhibits a few peculiarities worth describing. Variant *PMS2*:c.2186_2187del, predicted to cause a frameshift that alters the protein’s amino acid sequence beginning at codon 729 and leads to a premature stop codon 6 codons downstream, is primarily classified as VUS in ClinVar. It was found in homozygosity in this particular sample (parents’ genotyping was not available). The InSiGHT[15] database also classifies this variant as uncertain, noting that the variant is likely to originate from a pseudogene. However, this does not seem to be the case for our patient since the variant and TTT, rather than TTC, at codon 751 are contained in the same aligned reads, confirming that these reads come from the *PMS2* gene, not from *PMS2CL* (pseudogene). Additional considerations were taken into account in an attempt to ascertain the classification of this variant as pathogenic or benign. On the pathogenicity front, it is noteworthy that a slightly shorter truncated protein caused by c.2192T>G, NM_000535.7(*PMS2*):c.2192T>G, p.Leu731Ter, is classified as pathogenic at ClinVar by multiple submitters with no conflicts, and it aligns with ACMG guidelines. The resulting truncated protein would be very similar to that produced by c.2186_2187del, with both cases predicted to undergo nonsense-mediated decay (NMD). A relevant publication also supports this direction; considering that variant c.2186_2187del occurs in exon 13, within a repeated dinucleotide (CTCT), it has been described that variant c.2184_2185del was detected in Turcot syndrome-affected siblings as compound heterozygotes (R134X/2184delTC)[16]. This allelic data might suggest a variant detected in trans with a pathogenic variant for a recessive disorder (PM3). In this report, both siblings were compound heterozygotes (R134X/2184delTC), and 2184delTC was confirmed to be of maternal origin through the analysis of both parental DNAs. Considering the above, this variant could be classified as pathogenic per ACMG guidelines (PVS1, PM3, and PP5).

However, conflicting evidence may challenge this conclusion.

First*, PMS2*:c.2186_2187del would retain the DQHA(X)2E(X)4E motif found at the C-terminus of the protein encoded by this gene, forming part of the active site of the nuclease. Additionally, even though the premature stop codon location is predicted to induce the mechanism of nonsense-mediated decay (NMD), the transcripts could potentially be resistant to degradation[17]. In this case, the protein function could potentially remain unaffected. Second, its frequency is 2.81% in gnomAD (African), where the number of homozygotes is 14, and 1.33% in ABraOM[18] (based on whole-genome sequencing of 1,171 Brazilians in a census-based cohort); the recommended frequency threshold for *PMS2* is 0.05%. While these frequencies do not, in themselves, serve as stand-alone evidence for benignity, they warrant further scrutiny, particularly considering the absence of information on phenotype correlation from these databases.

Further evaluation regarding the consequence of *PMS2*:c.2186_2187del seems necessary, given the gene’s relevance to HBOC, hereditary nonpolyposis colon cancer, and constitutional mismatch repair deficiency syndrome. Functional studies might provide additional insights into this matter. In this small sample study, the prevalence of pathogenic mutations was somewhat higher than expected, at 25%. In a much larger evaluation with comparable patient inclusion criteria, albeit involving a different ethnic population and a considerably narrower gene panel, Shao et al.[19]found deleterious mutations at a rate of 19.50%.

## Materials and Methods

Patients were loosely selected based on a previous history of breast cancer at an early age and/or a first-degree/second-degree relative diagnosed with cancer (specific family relationship and cancer type information might be obtained by contacting the corresponding author), as shown in sTable 3. This project received approval from the Genoprimer Diagnostico Molecular Research Ethics Board (approval n° 011), and all individuals provided written consent for NGS testing.

**Table 3.** Basic characteristics of the study population.

| Patient ID | Age range at breast cancer diagnostic | Ethnicity<sup>a</sup> |
| --- | --- | --- |
| GCS003 | NA* | Central and Southern European |
| GCS005 | 41-45 | Iberian |
| GCS006 | 51-55 | Iberian |
| GCS007 | 41-45 | Indigenous |
| GCS008 | 51-55 | Southern European |
| GCS009 | 41-45 | Southern and Eastern European |
| GCS010 | 46-50 | Eastern European, Iberian |
| GCS011 | 41-45 | Indigenous |
| GCS012 | 26-30 | Iberian |
| GCS013 | 41-45 | Indigenous |
| GCS014 | 56-60 | Southern European |
| GCS015 | 36-40 | Central and Southern European |

NA: Not applicable, \*: No diagnosed cancer, ±: All Brazilian-born individuals descended from indigenous families or indicated region.

Genomic DNA was extracted from peripheral blood samples using standard methods. Gene panel library preparation was performed using three different kits: SureSelect XT HS2 DNA Reagent Kit®[20], QIAseq Targeted DNA Pro Human Hereditary Breast and Ovarian Cancer Panel - PHS-201Z-12, QIAGEN®[21], and Twist Target Enrichment Protocol[22], following the manufacturer’s instructions.

Pertinent data were generated in the standard order:

### Primary Analysis

FastQ files were obtained through next-generation sequencing performed on Illumina’s MiSeq System® with the Micro Kit v2 (300 cycles) flow cell.

### Secondary Analysis

Starting with paired-end reads, FastQ files underwent secondary analysis by aligning them to the GRCh38/UCSC hg38 genome using BWA[23]. Duplicate readings were removed, and variants (SNPs/indels) were detected with GATK HaplotypeCaller[24], generating VCF files.

### Tertiary Analysis

Variant annotation, filtering, and prioritization were carried out using “Franklin by Genoox[25],” enabling a straightforward analysis for SNPs/indels. Data visualization was achieved through Integrative Genomics Viewer (IGV)[26], which was crucial for reviewing data quality and reliability.

The gene panel was designed to encompass a comprehensive set of fifty genes that have been associated with HBOC, as shown in Table 4. The listed genes had their coding exon regions sequenced, extended to 10 bases from the 3’ end and 10 bases from the 5’ end.

**Table 4:** Genes associated with HBOC included in the panel.

|  |  |  |  |  |
| --- | --- | --- | --- | --- |
| *BRCA1* | *CHEK2* | *MRE11* | *SDHB* | *RINT1* |
| *BRCA2* | *ERBB2* | *NBN* | *SMARCA4* | *XRCC2* |
| *FGFR2* | *ESR1* | *STK11* | *TSHR* | *EPCAM* |
| *MAP3K1* | *MLH1* | *TP53* | *ABRAXAS1* | *HMMR* |
| *RAD51C* | *MSH2* | *RECQL* | *BLM* | *MUTYH* |
| *RAD51D* | *MSH6* | *XRCC3* | *DICER1* | *NQO2* |
| *TOX3* | *PALB2* | *NF1* | *FANCC* | *PGR* |
| *ATM* | *PIK3CA* | *PTEN* | *FANCM* | *PHB* |
| *BARD1* | *PMS2* | *APC* | *RAD50* | *POLD1* |
| *BRIP1* | *CDH1* | *AXIN2* | *RAD51* | *PRKAR1A* |

## Supporting information

S1 Table. Variants detected, including reference sequences.

Statement (resubmission)

## Data Availability

All data produced in the present study are available upon reasonable request to the authors.

## Acknowledgments

We extend our heartfelt appreciation to the dedicated individuals working in the clinical laboratory mentioned above, whose unwavering commitment to precision and excellence significantly contributed to the success of this research: Kamilla Leitão, Amabile Chacon, and Jessica Liaw.

## Supporting information

**S1 Table. Variants detected, including reference sequences.**

DP: Read Depth, Qual: Quality, AB: Allelic Balance, FS: Fisher Strand Bias, NA: Not Available.

## Notes

### Competing Interest Statement

The authors have declared no competing interest.

### Funding Statement

This study did not receive any funding

### Author Declarations

Genoprimer Diagnostico Molecular Research Ethics Board of Genoprimer Diagnostico Molecular gave ethical approval for this work.

