## Supplementary material for "Hereditary breast cancer next-generation sequencing (NGS) panel evaluation in the south region of Brazil: a novel *BRCA2* candidate pathogenic variant is reported": Statement (resubmission)

Dear The medRxiv team,

Statement

The IDs (e.g., GCS003, GCS015, etc. - as presented in Tables 1-3 and S1_Table) referenced in this study are unique identifiers created exclusively for the purpose of this paper publication. These identifiers are known only to the authors and were assigned specifically for use within the context of this research. They are not associated with any laboratory staff or patient records and consequently are not known to either patients or aboratory staff.

1. Mentions of precise ages have been replaced with age ranges.

2. Specific family relationship have been removed and advised that readers contact the corresponding author to request access to these data.

CESAR AUGUSTO BARROS DUARTE:78120349920

Digitally signed by CESAR AUGUSTO BARROS DUARTE:78120349920
Date: 2024.02.08 10:25:08 -03'00'
